# Engineering Analysis of Aortic Wall Stress in the Surgery of V-shape Resection of the Noncoronary Sinus

**DOI:** 10.1101/2020.12.02.20239475

**Authors:** Hai Dong, Minliang Liu, Tongran Qin, Liang Liang, Bulat Ziganshin, Hesham Ellauzi, Mohammad Zafar, Sophie Jang, John Elefteriades, Wei Sun

## Abstract

Ascending aortic aneurysms often include the sinotubular junction (STJ) and extend into the root portion of the aorta. The novel surgery of the V-shape resection of the noncoronary sinus of the aortic root has been shown to be a simpler procedure, comparing with traditional surgeries such as full aortic root replacement, for patients with moderate ascending aortic aneurysm and aortic root ectasia. This novel surgery could reduce the diameter and cross-sectional area of the aortic root. However, the detailed effect on the stress field and the rupture risk of the aortic root and aneurysm has not been fully investigated. In this study, we performed patient-specific finite element (FE) analysis based on the 3D geometries of the aortic root and ascending aortic aneurysm, reconstructed directly from the clinical computed tomographic (CT) images. By comparing the pre- and post-surgery results, we investigated the influence of the V-shape surgery on the stress field and rupture risk of the aortic root, ascending aortic aneurysm and aortic arch. It was found that the surgery could significantly reduce the wall stress of the aortic root, ascending aortic aneurysm, as well the aortic arch, and hence lower the rupture risk.

## 1. Introduction

Aortic aneurysm often claims lives without any premonitory symptoms or signs, and is a disease among the leading causes of death of adults [1, 2]. This disease shows notable clinical risk and about half of untreated aortic aneurysms in high-risk patients could rupture within 1 year [3, 4]. Ascending aortic aneurysms (AsAA) often include the dilatation of sinotubular junction (STJ) and extend into the root part of the aorta, but usually influence the noncoronary or right coronary sinus [5]. The pathology of AsAA with root ectasia often leads to aortic insufficiency. The pathology of the root, valve and ascending aorta can be eradicated by traditional surgeries such as full aortic root replacement. However, this kind of surgery may represent excessive surgical intervention for infirm or elderly patients. The novel surgery [6] of the V-shape resection of the noncoronary sinus has been shown to be a simpler procedure, comparing with the traditional surgery of full root replacement. The novel surgery could reduce the diameter and cross-sectional area of the aortic aneurysm [6]. However, its detailed effect on the stress field and the rupture risk of the aortic root and ascending aortic aneurysm has not been fully investigated.

It usually requires the in vivo material parameters of the aortic tissue to calculate the patient-specific stress field of aorta by conventional approaches [7-11]. However, to obtain the in vivo material parameters [12-16] usually needs multi-phase images of the aorta which may not be available all the time. Fortunately, recent studies [15, 17] have shown that the aorta is approximately statically determinate and the wall stress is nearly insensitive to the variation of material properties. Then, the aortic wall stress can be calculated by the static determinacy approach based on only one phase CT image without knowing the in vivo material parameters. The static determinacy approach is usually based on thin-walled models [17, 18] which are incompatible with the residual deformation. Thus it is usually considered as a limitation that the residual deformation/stress is not considered by the static determinacy approach. However, more recently, the study by our group [19] showed that the residual deformation/stress may be not a limitation of the static determinacy approach by demonstrating that the transmural mean stress of aortic wall is approximately the same for the thin-walled models and thick-walled models. Thus, the transmural mean stress of the aortic wall can be readily obtained from in vivo clinical images without knowing the patient-specific material properties and residual deformations.

In this study, we performed finite element (FE) analysis, based on the static determinacy approach, of the aorta pre- and post-surgery of patients who underwent the novel surgery of V-shape resection of the noncoronary sinus. The 3D geometries of the aorta including the aortic root, ascending aorta, aortic arch, and descending aorta, used in the FE analysis, were reconstructed from the clinical computed tomographic (CT) images. We obtained the aortic wall stress and investigated the influence of the V-shape resection of the noncoronary sinus on the stress field and rupture risk of the aortic root, ascending aortic aneurysm and aortic arch.

## 2. Materials and methods

### 2.1. Patient Data and the V-shape surgery

Deidentified clinical cardiac CT scans were obtained for a total of 10 patients who underwent the surgery of V-shape noncoronary sinus resection, together with supracoronary ascending aortic replacement, at Yale-New Haven Hospital between the years of 2015 and 2018. During the surgery, the aorta is circularly opened just above the STJ. Then a deep V-shaped or triangular portion of the noncoronary sinus of the aortic root is resected and the root wall is directly reapproximated in two layers, one everting mattress suture layer and another running over-and-over layer. The detailed procedure of the V-shape surgery can be found in Ref. [6]. All human subjects in this study provided written and informed consent, and IRB-approved protocols were obtained from Georgia Tech Institutional Review Board and Yale University Institutional Review Board. A set of approximately 300–2000 axial CT images containing the thoracic and abdominal aorta pre- and post-surgery, with a resolution of 1.2 mm by 1.2 mm by 1.0 mm, were obtained for each patient.

### 2.2 Construction of Patient-Specific Finite Element Model

For each patient, the three-dimensional surface geometry of the aorta was reconstructed from the CT images using 3D Slicer (www.slicer.org). The aorta was segmented in 3D Slicer with the module of Robust Statistics Segmenter, following painting seeds in the Editing module. The aortic root, ascending aorta, aortic arch together with the three branches around it, and the descending aorta were obtained (Fig. 1a). The surface geometry was exported from 3D Slicer and imported into Altair HyperMesh 2017 (Altair Engineering) to generate the finite elements. We first created 4-node quadrilateral shell elements (S4R) with elements size of about 2 mm by 2mm based on the surface geometry. Four layers of 8-node linear brick elements (C3D8R) were created by offsetting the S4R elements with a total thickness of 1.5 mm (Fig. 1b). Mesh independence analyses were performed.

**Fig 1.**
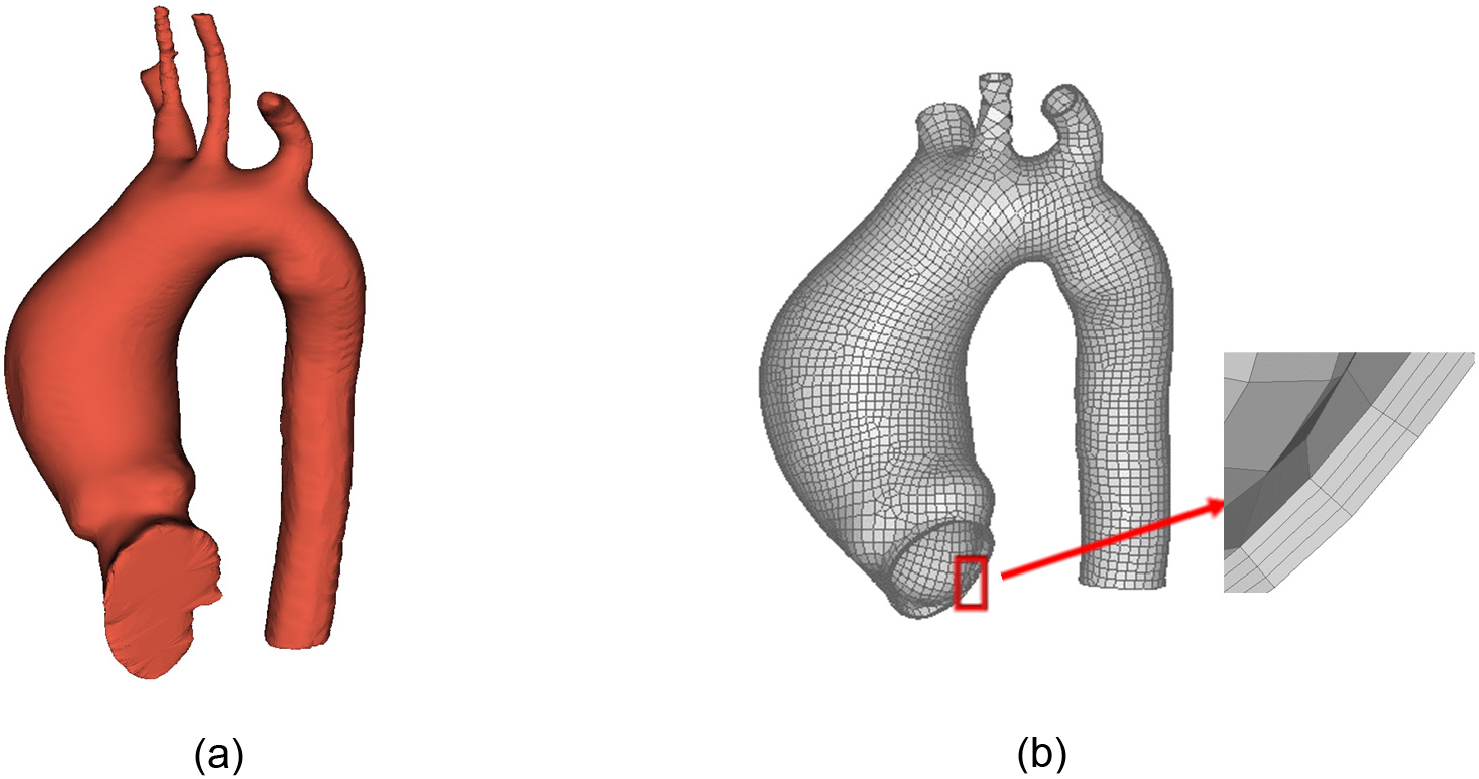
(a) 3D geometry of the aorta of a representative patient pre-surgery reconstructed in 3D Slicer; (b) Finite element model generated in HyperMesh based on the geometry in (a) with the trimmed boundaries.

### 2.3 Finite element analyses based on static determinacy approach

Using the Laplace law to compute the wall hoop stress of a perfect cylindrical tube is one of the well-known examples of static determinacy [19], of which the wall hoop stress *σ*_*θθ*_ can be expressed as

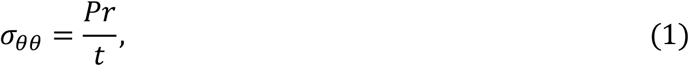

where *P* is the inner pressure, *r* is the radius of the tube and *t* is the wall thickness of the tube. Eq. (1) shows that the wall hoop stress of the tube is independent of material properties and can be directly determined from the static force balance.

Though the geometry of aorta is much more complex than a perfect cylindrical tube, it has been shown [15, 17, 19] that the aorta is statically determinate and the wall stress is insensitive to the variation of material properties. We performed finite element (FE) simulation of the aorta by the Abaqus/Standard 2019 (SIMULIA, Providence, RI) based on the static determinacy approach. During the FE simulation, isotropic material properties with extremely large stiffness were applied to the aortic wall, with the Young’s modulus to be *E* = 20 GPa and the Poisson’s ratio to be *v* = 0.49. The circumferential and axial displacements were fixed for all the boundaries, including the inlet of the ascending aorta, outlet of the three branches, and outlet of the descending aorta. The radial displacement of the boundaries was set to be free. A systolic pressure of 120 mmHg was applied to the inner surface of the aorta during the simulation. The stress field of the aorta pre- and post-surgery was extracted and analyzed.

## 3. Results

The wall stress of the aorta of 10 patients (P1-P10) pre- and post-surgery was obtained. Fig. 2 shows the max-principal (MP) stress field of the aorta of a representative patient (P1) pre- and post-surgery. We extracted the MP stress field of four ring-bands of the aorta of each patient, with band-1 at the aortic root, band-2 at the ascending aneurysm position with the maximum diameter pre-surgery, band-3 at the ascending aneurysm position with diameter un-affected by the surgery and band-4 at the position right distal to the aortic arch. The ascending aneurysm of band-2 pre-surgery was replaced by an ascending aortic graft post-surgery. Band-1, band-3 and band-4 post-surgery are live aortic tissue. The mean and peak MP stress of the four ring bands of the 10 patients were presented in Table 1. The green background in Table 1 represents a significant decrease (absolute value greater than 5.0%) of the stress from pre- to post-surgery, blue background represents little change (absolute value less than 5.0%) of the stress, and orange background represents a significant increase (greater than 5.0%) of the stress.

**Table 1:** Mean and peak MP stress of the four ring bands (Fig. 2) of the 10 patients. Pre: stress of the band of a patient pre-surgery; Post: stress of the band of a patient post-surgery; Chg: percentage change of the stress from pre- to post-surgery. Green background represents a significant decrease of the stress from pre- to post-surgery, blue background represents little change (<5.0%) of the stress, and orange background represents a significant increase of the stress.

|  | Band-1 |  | Band-2 |  | Band-3 |  | Band-4 |  |
| --- | --- | --- | --- | --- | --- | --- | --- | --- |
| (kPa) | Mean | Peak | Mean | Peak | Mean | Peak | Mean | Peak |
| P1-Pre | 226.9 | 321.9 | 233.8 | 400.7 | 219.3 | 263.3 | 160.0 | 303.3 |
| P1-Post | 162.2 | 244.6 | 137.5 | 169.4 | 158.4 | 232.7 | 139.6 | 212.9 |
| P1-Chg | -28.5% | -24.0% | -41.2% | -57.7% | -27.8% | -11.6% | -12.8% | -29.8% |
| P2-Pre | 241.9 | 420.8 | 237.3 | 426.8 | 233.3 | 328.3 | 228.1 | 487.0 |
| P2-Post | 168.1 | 296.6 | 146.0 | 226.9 | 182.2 | 290.9 | 159.8 | 209.0 |
| P2-Chg | -30.5% | -29.5% | -38.5% | -46.8% | -21.9% | -11.4% | -29.9% | -57.1% |
| P3-Pre | 242.7 | 401.6 | 250.4 | 371.8 | 226.5 | 287.3 | 169.5 | 282.6 |
| P3-Post | 200.0 | 349.2 | 179.2 | 286.3 | 196.1 | 264.0 | 166.4 | 243.7 |
| P3-Chg | -17.6% | -13.0% | -28.4% | -23.0% | -13.4% | -8.1% | -1.8% | -13.8% |
| P4-Pre | 245.4 | 539.5 | 247.8 | 379.9 | 249.5 | 342.6 | 169.3 | 300.7 |
| P4-Post | 193.5 | 344.2 | 163.5 | 244.1 | 207.8 | 282.2 | 173.7 | 286.1 |
| P4-Chg | -21.1% | -36.2% | -34.0% | -35.7% | -16.7% | -17.6% | 2.6% | -4.9% |
| P5-Pre | 259.7 | 384.9 | 265.1 | 380.4 | 247.1 | 317.3 | 169.6 | 237.9 |
| P5-Post | 177.9 | 310.0 | 167.0 | 246.6 | 196.3 | 278.8 | 151.4 | 232.4 |
| P5-Chg | -31.5% | -19.5% | -37.0% | -35.2% | -20.6% | -12.1% | -10.7% | -2.3% |
| P6-Pre | 265.3 | 911.4 | 287.4 | 493.9 | 282.1 | 438.3 | 207.1 | 360.3 |
| P6-Post | 182.4 | 438.5 | 161.6 | 264.4 | 202.6 | 420.2 | 153.1 | 208.5 |
| P6-Chg | -31.2% | -51.9% | -43.8% | -46.5% | -28.2% | -4.1% | -26.1% | -42.1% |
| P7-Pre | 212.6 | 382.3 | 243.9 | 400.1 | 220.4 | 275.9 | 152.9 | 277.6 |
| P7-Post | 177.9 | 310.0 | 167.0 | 246.6 | 196.3 | 278.8 | 151.4 | 232.4 |
| P7-Chg | -16.3% | -18.9% | -31.5% | -38.4% | -10.9% | 1.1% | -1.0% | -16.3% |
| P8-Pre | 242.7 | 387.9 | 259.6 | 520.7 | 232.3 | 347.6 | 192.1 | 353.5 |
| P8-Post | 184.5 | 297.5 | 191.9 | 332.2 | 193.1 | 349.1 | 194.1 | 361.8 |
| P8-Chg | -24.0% | -23.3% | -26.1% | -36.2% | -16.9% | 0.4% | 1.0% | 2.3% |
| P9-Pre | 252.8 | 564.8 | 225.9 | 395.3 | - | - | 163.6 | 265.7 |
| P9-Post | 188.4 | 308.7 | 151.1 | 169.6 | - | - | 170.1 | 409.0 |
| P9-Chg | -25.5% | -45.3% | -33.1% | -57.1% | - | - | 4.0% | 53.9% |
| P10-Pre | 227.0 | 388.1 | 234.0 | 429.2 | 209.5 | 303.8 | 153.4 | 202.6 |
| P10-Post | 166.5 | 269.5 | 221.4 | 324.0 | 212.2 | 351.8 | 164.2 | 230.1 |
| P10-Chg | -26.7% | -30.6% | -5.4% | -24.5% | 1.3% | 15.8% | 7.0% | 13.6% |

**Fig 2.**
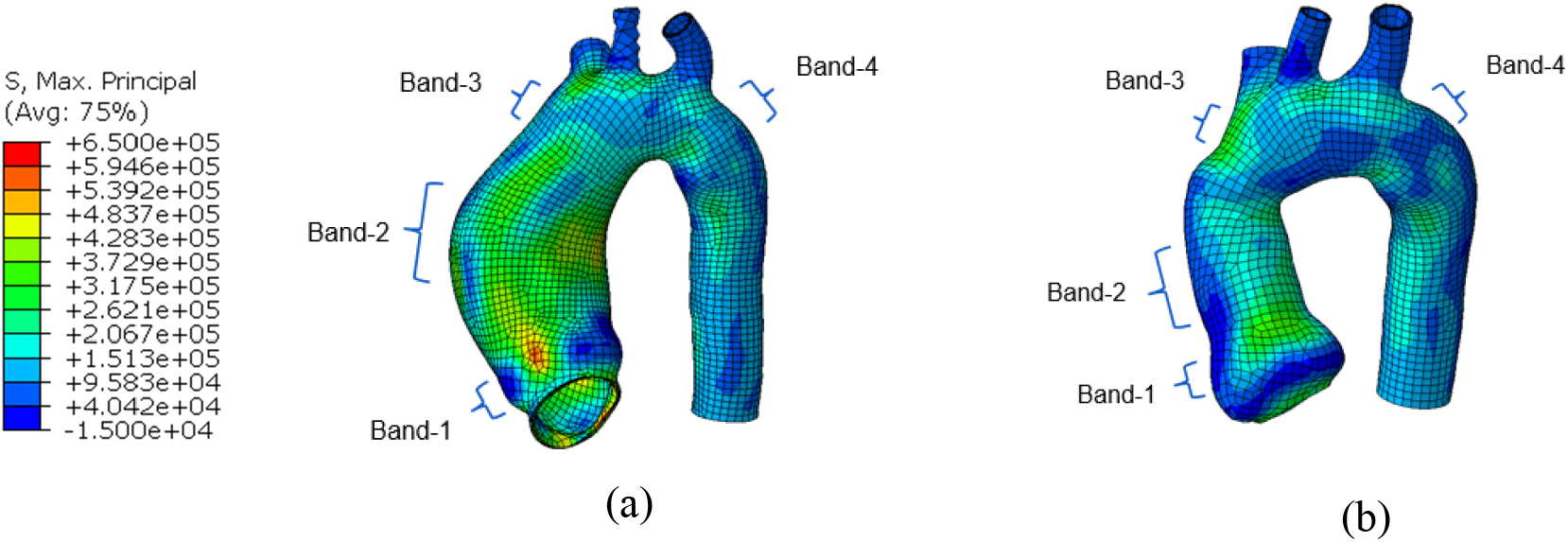
Max-principal stress field of the aorta of a representative patient (P1): (a) pre- and (b) post-surgery. Four ring bands: band-1 at the aortic root, band-2 at the ascending aneurysm position with the maximum diameter pre-surgery, band-3 at the ascending aneurysm position with diameter un-affected by the surgery and band-4 at the position right distal to the aortic arch.

The results show that the mean and peak MP stress of the aortic root (band-1) of all the 10 patients were reduced significantly by the novel V-shape surgery, with a maximum decrease of −31.5% (P5) and a minimum decrease of −16.3% (P7) for the mean MP stress. The maximum decrease of the peak MP stress of the root reaches −51.9% (P6) and the minimum decrease of the peak MP stress of the root is −13.0% (P3). For the ascending aneurysm position (band-2) with the maximum diameter pre-surgery, the maximum and minimum decrease of the mean MP stress are −43.8% (P6) and −5.4% (P10), respectively. The maximum and minimum decrease of the peak MP stress of band-2 are −57.7% (P1) and −23.0% (P3), respectively.

For the ascending aneurysm position with diameter un-affected by the surgery (band-3), the mean and peak MP stress of P1-P5 are significantly decreased by the surgery, with the maximum decrease of the mean and peak MP stress reaches −27.8% (P1) and −17.6% (P4). For P6, P7 and P8, the mean MP stress of band-3 decreases significantly post-surgery but the peak MP stress does not change much (less than 4.5%). For P9, the stress of band-3 was not obtained since most of the ascending aorta has been replaced by the ascending aortic graft post-surgery (see Fig. 3). For P10, the mean MP stress of band-3 changes little (less than 2%) but the peak MP stress has an obvious increase post-surgery.

**Fig 3.**
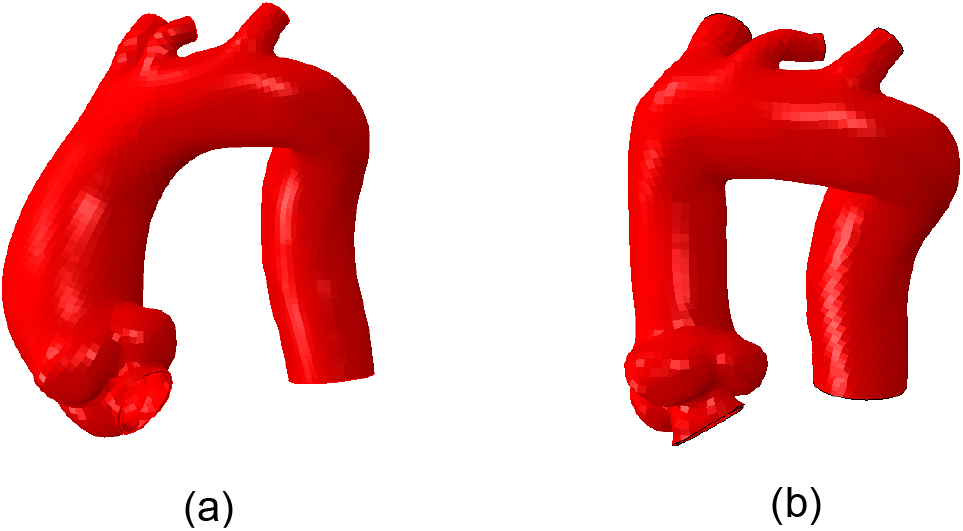
Pre-surgery (a) and post-surgery (b) geometries of the aorta of P9. Most of the ascending aorta has been replaced by the ascending aortic graft and thus the stress of band-3 as shown in Fig. 1 was not obtained for P9.

For the position right distal to the aortic arch (band-4 in Fig. 2), the mean MP stress decreases for P1, P2, P5 and P6, with the maximum decrease reaching −29.9% (P2). The mean MP stress of band-4 changes little (less than 4%) for P3, P4, and P7-P9. However, the mean MP stress of band-4 for P10 exhibits an increase of 7.0%. The peak MP stress of band-4 significantly decreases for P1-P3, P6, and P7, but changes not much for P4, P5, and P8 (less than 5%). The peak MP stress of band-4 exhibits an obvious increase for P9 and P10.

## 4. Discussions

The V-shape resection of the noncoronary sinus and remodeling of the aortic root reduce the diameter of the root, and thus reduces the wall stress of the root for all 10 patients in Table 1. If the rupture strength of the wall is assumed to be unchanged post-surgery, the rupture risk will decrease with the decreasing wall stress [20-22]. The major ascending aneurysm (band-2 in Fig. 2) was replaced by an ascending aortic graft. The diameter of the graft is much smaller than that of the ascending aneurysm pre-surgery and thus the stress of band-2 also reduces for all the 10 patients. Although the tough graft post-surgery has no rupture issue, the reduction of the diameter of band-2 may contribute to the stress reduction of the adjacent band-3 (live aortic tissue) for most patients in Table 1.

It should be noted that there is an increase for the peak MP stress of band-4 of P9, the peak stress of band-3 of P10, and the mean and peak stress of band-4 of P10. Studies in the literature [23-25] show that the wall stress of the aorta was affected by the overall curvature and tortuosity of the aorta. The potential reason for the stress increase of band-3 and band-4 of P9 and P10 could be the change of the curvature, tortuosity and overall geometry of the aorta post-surgery (see Fig. 2 for P9). In the future work, the detailed dependence of the stress change on the overall geometry change will be investigated.

One limitation of this study is that only the transmural mean stress of the aortic wall was investigated and the stress variation in the wall thickness direction was not considered, because the through-thickness distribution of the stress may not be obtained accurately based on the static determinacy approach [19]. Another limitation is that a uniform thickness of 1.5 mm for the aortic wall was applied in the FE simulation, while the thickness of the aortic wall may be non-uniform. In the future, in vivo non-uniform thickness of the aortic wall may be obtained based on the method in Ref. [26] when the contrast and non-contrast CT scans are available.

## 5. Conclusions

In this study, we reconstructed the patient-specific 3D geometries of aorta pre- and post-surgery, from the clinical computed tomographic (CT) images, of the patients who underwent the novel surgery of V-shape resection of the noncoronary sinus. We performed finite element (FE) analysis of the aorta, including the aortic root, ascending aorta, aortic arch, and descending aorta, based on the static determinacy approach. We obtained the aortic wall stress of the ascending aortic aneurysm pre- and post-surgery, and investigated the influence of the V-shaped resection of the noncoronary sinus on the stress field and rupture risk of the aortic root, ascending aortic aneurysm and aortic arch. The results show that the V-shape surgery could significantly reduce the wall stress of the aortic root, ascending aortic aneurysm, as well the aortic arch, and hence lower the rupture risk.

## Data Availability

The data have been presented in the manuscript and there are no external data

## 6. Acknowledgment

We thank the lab members Kristen Riess, Courtney Huynh, Samuel Li, Parikirt Oggu and Daksha Jadhav for assistance in image segmentation, mesh creation and finite element simulation.

## References

[1] CDC, 2007, “National Center for Injury Prevention and Control, WISQARS Leading Causes of Death Reports, 1999–2007”.

[2] C. Martin, W. Sun, T. Pham, J. Elefteriades, Predictive biomechanical analysis of ascending aortic aneurysm rupture potential, Acta biomaterialia 9(12) (2013) 9392–9400.

[3] J. Humphrey, C. Taylor, Intracranial and abdominal aortic aneurysms: similarities, differences, and need for a new class of computational models, Annu. Rev. Biomed. Eng. 10 (2008) 221–246.

[4] C. Martin, W. Sun, J. Elefteriades, Patient-specific finite element analysis of ascending aorta aneurysms, American Journal of Physiology-Heart and Circulatory Physiology 308(10) (2015) H1306–H1316.

[5] S. Westaby, S. Saito, K. Anastasiadis, N. Moorjani, X. Jin, Aortic root remodeling in atheromatous aneurysms: the role of selected sinus repair, European journal of cardio-thoracic surgery 21(3) (2002) 459–464.

[6] J.A. Elefteriades, S. Peterss, N. Nezami, G. Gluck, W. Sun, M. Tranquilli, B.A. Ziganshin, V-shape noncoronary sinus remodeling in ascending aortic aneurysm and aortic root ectasia, The Journal of thoracic and cardiovascular surgery 154(1) (2017) 72–76.

[7] V. Alastrué, A. Garía, E. Peña, J. Rodríguez, M. Martínez, M. Doblaré, Numerical framework for patient-specific computational modelling of vascular tissue, International journal for numerical methods in biomedical engineering 26(1) (2010) 35–51.

[8] D.M. Pierce, T.E. Fastl, B. Rodriguez-Vila, P. Verbrugghe, I. Fourneau, G. Maleux, P. Herijgers, E.J. Gomez, G.A. Holzapfel, A method for incorporating three-dimensional residual stretches/stresses into patient-specific finite element simulations of arteries, Journal of the mechanical behavior of biomedical materials 47 (2015) 147–164.

[9] S. Ruiz de Galarreta, A. Cazón, R. Antón, E.A. Finol, A methodology for verifying abdominal aortic aneurysm wall stress, Journal of biomechanical engineering 139(1) (2017).

[10] S.J. Mousavi, S. Farzaneh, S. Avril, Computational predictions of damage propagation preceding dissection of ascending thoracic aortic aneurysms, International journal for numerical methods in biomedical engineering 34(4) (2018) e2944.

[11] T.C. Gasser, M. Auer, F. Labruto, J. Swedenborg, J. Roy, Biomechanical rupture risk assessment of abdominal aortic aneurysms: model complexity versus predictability of finite element simulations, European Journal of Vascular and Endovascular Surgery 40(2) (2010) 176–185.

[12] M. Kroon, G.A. Holzapfel, Elastic properties of anisotropic vascular membranes examined by inverse analysis, Computer Methods in Applied Mechanics and Engineering 198(45-46) (2009) 3622–3632.

[13] J. Lu, X. Zhao, Pointwise identification of elastic properties in nonlinear hyperelastic membranes— part I: theoretical and computational developments, Journal of applied mechanics 76(6) (2009).

[14] X. Zhao, X. Chen, J. Lu, Pointwise identification of elastic properties in nonlinear hyperelastic membranes—part II: experimental validation, Journal of applied mechanics 76(6) (2009).

[15] M. Liu, L. Liang, W. Sun, A new inverse method for estimation of in vivo mechanical properties of the aortic wall, Journal of the mechanical behavior of biomedical materials 72 (2017) 148–158.

[16] L. Liang, M. Liu, C. Martin, W. Sun, A machine learning approach as a surrogate of finite element analysis-based inverse method to estimate the zero-pressure geometry of human thoracic aorta, International journal for numerical methods in biomedical engineering (2018) e3103.

[17] G.R. Joldes, K. Miller, A. Wittek, B. Doyle, A simple, effective and clinically applicable method to compute abdominal aortic aneurysm wall stress, Journal of the mechanical behavior of biomedical materials 58 (2016) 139–148.

[18] J. Lu, Y. Luo, Solving membrane stress on deformed configuration using inverse elastostatic and forward penalty methods, Computer Methods in Applied Mechanics and Engineering 308 (2016) 134–150.

[19] M. Liu, L. Liang, H. Liu, M. Zhang, C. Martin, W. Sun, On the computation of in vivo transmural mean stress of patient-specific aortic wall, Biomech Model Mechanobiol 18(2) (2019) 387–398.

[20] H. Dong, J. Wang, B.L. Karihaloo, An improved Puck’s failure theory for fibre-reinforced composite laminates including the in situ strength effect, Composites science and technology 98 (2014) 86–92.

[21] H. Dong, J. Wang, A criterion for failure mode prediction of angle-ply composite laminates under in-plane tension, Composite Structures 128 (2015) 234–240.

[22] M. Liu, H. Dong, X. Lou, G. Iannucci, E.P. Chen, B.G. Leshnower, W. Sun, A Novel Anisotropic Failure Criterion With Dispersed Fiber Orientations for Aortic Tissues, Journal of Biomechanical Engineering 142(11) (2020).

[23] L. Liang, M. Liu, C. Martin, J.A. Elefteriades, W. Sun, A machine learning approach to investigate the relationship between shape features and numerically predicted risk of ascending aortic aneurysm, Biomech Model Mechanobiol (2017).

[24] S. Pappu, A. Dardik, H. Tagare, R.J. Gusberg, Beyond fusiform and saccular: a novel quantitative tortuosity index may help classify aneurysm shape and predict aneurysm rupture potential, Annals of vascular surgery 22(1) (2008) 88–97.

[25] M. Piccinelli, S. Bacigaluppi, E. Boccardi, B. Ene-Iordache, A. Remuzzi, A. Veneziani, L. Antiga, Geometry of the internal carotid artery and recurrent patterns in location, orientation, and rupture status of lateral aneurysms: an image-based computational study, Neurosurgery 68(5) (2011) 1270–1285.

[26] J.A. Elefteriades, S.K. Mukherjee, H. Mojibian, Discrepancies in Measurement of the Thoracic Aorta: JACC Review Topic of the Week, Journal of the American College of Cardiology 76(2) (2020) 201–217.

